# COVID-19 incidence and R decreased on the Isle of Wight after the launch of the Test, Trace, Isolate programme

**DOI:** 10.1101/2020.07.12.20151753

**Authors:** Michelle Kendall, Luke Milsom, Lucie Abeler-Dörner, Chris Wymant, Luca Ferretti, Mark Briers, Chris Holmes, David Bonsall, Johannes Abeler, Christophe Fraser

## Abstract

In May 2020 the UK introduced a Test, Trace, Isolate programme in response to the COVID-19 pandemic. The programme was first rolled out on the Isle of Wight and included Version 1 of the NHS contact tracing app. We used COVID-19 daily case data to infer incidence of new infections and estimate the reproduction number R for each of 150 Upper Tier Local Authorities in England, and at the National level, before and after the launch of the programme on the Isle of Wight. We used Bayesian and Maximum-Likelihood methods to estimate R, and compared the Isle of Wight to other areas using a synthetic control method. We observed significant decreases in incidence and R on the Isle of Wight immediately after the launch. These results are robust across each of our approaches. Our results show that the sub-epidemic on the Isle of Wight was controlled significantly more effectively than the sub-epidemics of most other Upper Tier Local Authorities, changing from having the third highest reproduction number R (of 150) before the intervention to the tenth lowest afterwards. The data is not yet available to establish a causal link. However, the findings highlight the need for further research to determine the causes of this reduction, as these might translate into local and national non-pharmaceutical intervention strategies in the period before a treatment or vaccination becomes available.

## Introduction

As part of efforts to control the spread of COVID-19 using non-pharmaceutical interventions, many countries are prioritising community testing, case isolation, contact tracing, and quarantining of contacts of cases [3]. Contact tracing usually requires individually questioning index cases (individuals identified as having COVID-19), relying on their memory of past close proximity contact events [4]; contact tracing may also be conducted via smartphone contact tracing apps which can be used to pass anonymised notifications between new index cases and their past contacts [5,6]. These two approaches are complementary: the former approach relies on noticing and remembering contacts and an efficient infrastructure of trained public health officials; the latter relies on establishing a well-calibrated new technology and appreciable population uptake and adherence. The former is a more familiar intervention and a phone call may be more effective once received, but the speed of notification of the latter may be critical given the speed of COVID-19 transmission [5]. In May 2020, a pilot of this combined approach was conducted on the Isle of Wight, an island to the south of England with 141,536 inhabitants, of whom 24% are over the age of 65 [7].

The UK national policy of “Test, Trace, Isolate” (TTI) included community testing and contact tracing conducted by Public Health England using the Contact Tracing and Advisory Service (CTAS). On the Isle of Wight it also included version 1 of the NHS contact tracing app. The app was configured for people to report two characteristic symptoms of COVID-19: cough and fever. Shortly after reporting symptoms, the app anonymously notified other app users who had been in recent contact with the individual, and offered generic guidance on how to reduce risks of transmission to others (though no instructions to self-isolate or quarantine).

The Isle of Wight TTI programme was launched on 5 May 2020, with the app made available for download for the general public from 7 May. The app was downloaded by more than 54,000 people on the island during the trial period, corresponding to 38% of the population [8]. On 18 May community testing was introduced nationwide, initially for all people above the age of 5, then on 28 May for everyone. Also on 28 May, manual contact tracing through the national contact tracing service was re-introduced nationwide. At the time of writing, there is no contact tracing app nationwide. The full timeline is shown in Figure 1.

**Figure 1:**
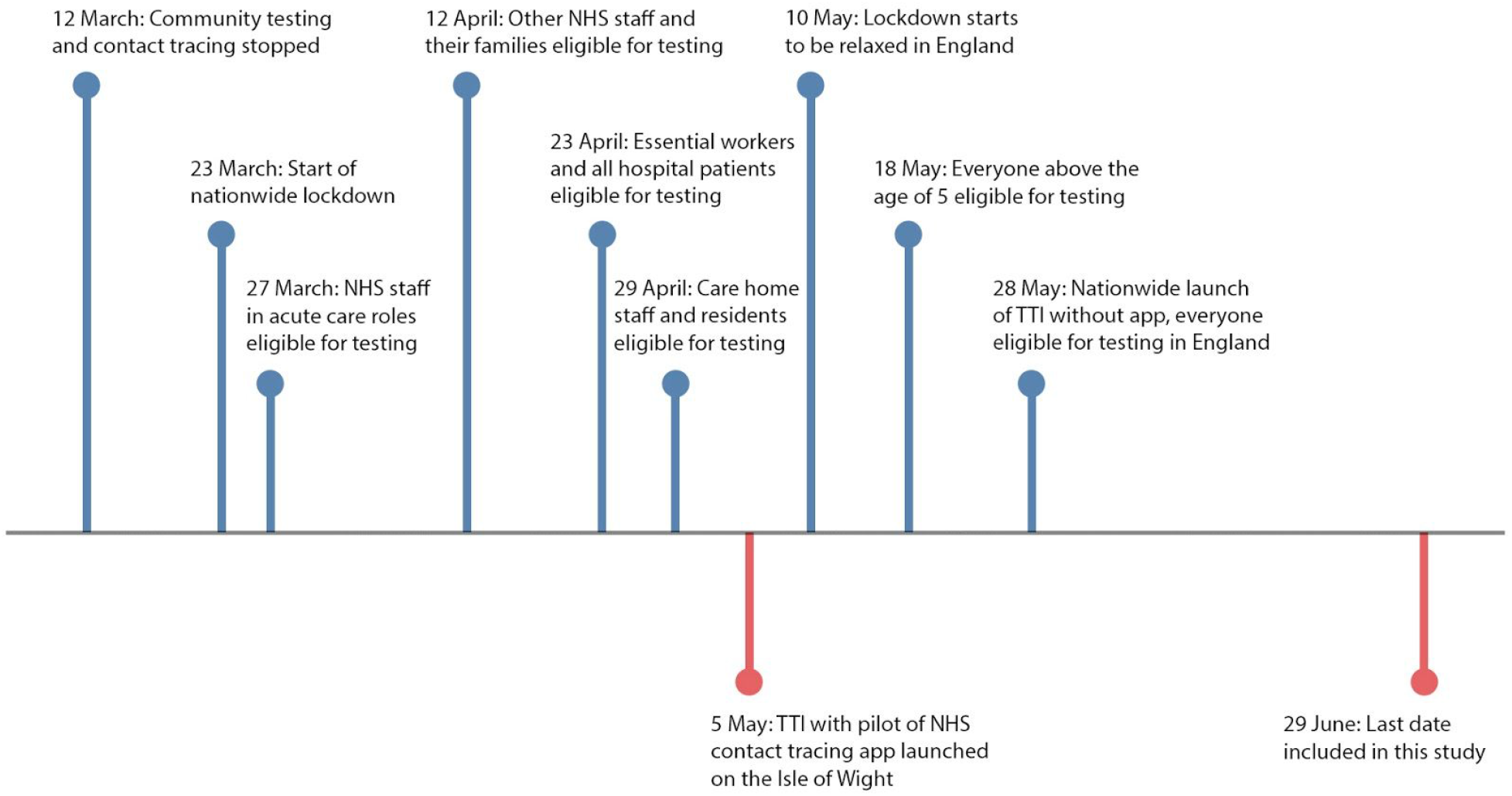
Timeline of events related to contact tracing and testing in England (blue) and the Isle of Wight (red).

Between the 6th May and 26 May, 160 cases on the Isle of Wight were reported to manual contact tracing, resulting in 163 individuals receiving a notification and request to self-isolate, and during the same period 1,524 people reported symptoms to the app resulting in 1,188 receiving an exposure notification.

In this study we report trends in incidence on the Isle of Wight before and during the period of this pilot, and compare these trends with other areas of England. We focus on total incidence, per capita incidence, and the reproduction number R. We do not analyse data on the function of the contact tracing intervention; we rather explore whether the intervention might have been associated with declines in rates of new infection that stand out compared to trends in other areas of England.

## Material and Methods

More information on data and methods can be found in the supplement.

### Data on incidence of confirmed cases by area

New laboratory-confirmed COVID-19 cases are reported daily for each of 150 areas of England, specifically Upper-tier Local Authorities (UTLAs), of which the Isle of Wight is one. There are two categories of case data: “Pillar 1” data which consists of tests conducted in hospitals, for those with a clinical need and health and care workers, and “Pillar 2” data from tests in the wider population [9]. Pillar 1 and Pillar 2 data represent tests taken at different stages of individuals’ disease progression, which needs to be considered for analysis. Estimates of the total population size for each UTLA were obtained from the Office for National Statistics [10].

### Back-calculation of infection times

We used a back-calculation approach to estimate the distribution of the time of infection in Pillar 1 data, assuming that most Pillar 1 tests are derived from hospital patients. We used a lognormal distribution with mean 5.42 days and standard deviation 2.7 days for the incubation period following a meta-analysis [11], and a gamma distribution with mean 5.14 days and standard deviation 4.2 days for the time from symptom onset to hospitalisation [12]. Analyses were truncated at 14 June as calculation beyond this date would have been affected by missing data 25-29 June [13].

### Estimate of the generation time

The generation time distribution is the probability distribution of times elapsed between an individual becoming infected and their transmitting to someone else. We used a gamma distribution with mean 5.5 days and standard deviation 2.14 days as obtained by Ferretti et al. (in prep.) combining data from [5, 14, 15, 16]. This distribution was converted into a discrete distribution on integer time intervals, i.e. 0,1,2,3,… days post infection.

### Renewal equation estimation for R(t)

We used Bayesian estimation of the instantaneous reproduction number [17] as implemented in the software package EpiEstim [18,19]. To reduce oversensitivity to the priors when the number of infections is very low, we used the posterior mode rather than the posterior mean as the central estimate.

### Nowcasting

We combined R with incidence per capita to provide a simple “nowcast” for each UTLA. For each day we multiplied the estimated R value by the mean incidence per capita for the week beginning three days before and ending three days afterwards.

### Creating synthetic controls for the 150 UTLAs in England

We used a ‘synthetic control’ approach [20,21] to construct a comparison area for the Isle of Wight. The approach creates a weighted average of other areas in England that matches the Isle of Wight in the average R before the introduction of the pilot, as well as in distributions of age and ethnicity. To choose the weights, we used a cross validation with a 19-day training period followed by a 19-day validation period. Significance was determined using a permutation test.

### Maximum-Likelihood estimation of R

Pillar 2 daily case data based on community testing was not publicly available at the time of writing for individual UTLAs, and the combined dataset available presents challenges for accurate inference of incidence of new infections, so we did not perform the Bayesian EpiEstim analysis on these datasets. Instead we used a Maximum-Likelihood estimation of the growth rate *r* over defined time periods which clearly post-date the TTI interventions, and from these estimated an average value of R for the Isle of Wight and at a national level [22]. We used publicly available data for the Pillar 1 and combined pillars comparisons, and for Pillar 2 we made use of Isle of Wight data obtained via private communication with Public Health England. We also used Pillar 1 data for each UTLA to compare values of R in time periods before and after the TTI interventions. This simple change-point analysis acts as a robustness check for our earlier analyses as it does not depend on the same modelling assumptions.

## Results

### The incidence of new COVID cases decreased both in the Isle of Wight and nationally during the period of study

In Figure 2 we show the number of new confirmed daily cases on the Isle of Wight and “nationally”, by individual pillars (Figures 2A and 2B) and using the combined pillars data released on 2 July 2020 (Figures 2C and 2D). By “nationally” we refer to the data available: England for Pillar 1, the “UK except Wales” for Pillar 2, and England for the combined pillars data. Pillar 1 data is available for the other nations of the UK over the same time period but suffered from sporadic reporting which made incidence and R difficult to estimate with confidence. We note that although it has become common to see Pillar 1 and 2 data displayed in stacked histograms such as these, interpreting true incidence from these plots is non-trivial because of the different lag times on testing infected individuals in the two pillars. Nevertheless, there is a clear trend of declining incidence through May and June, even in the context of increasing availability of tests.

**Figure 2:**
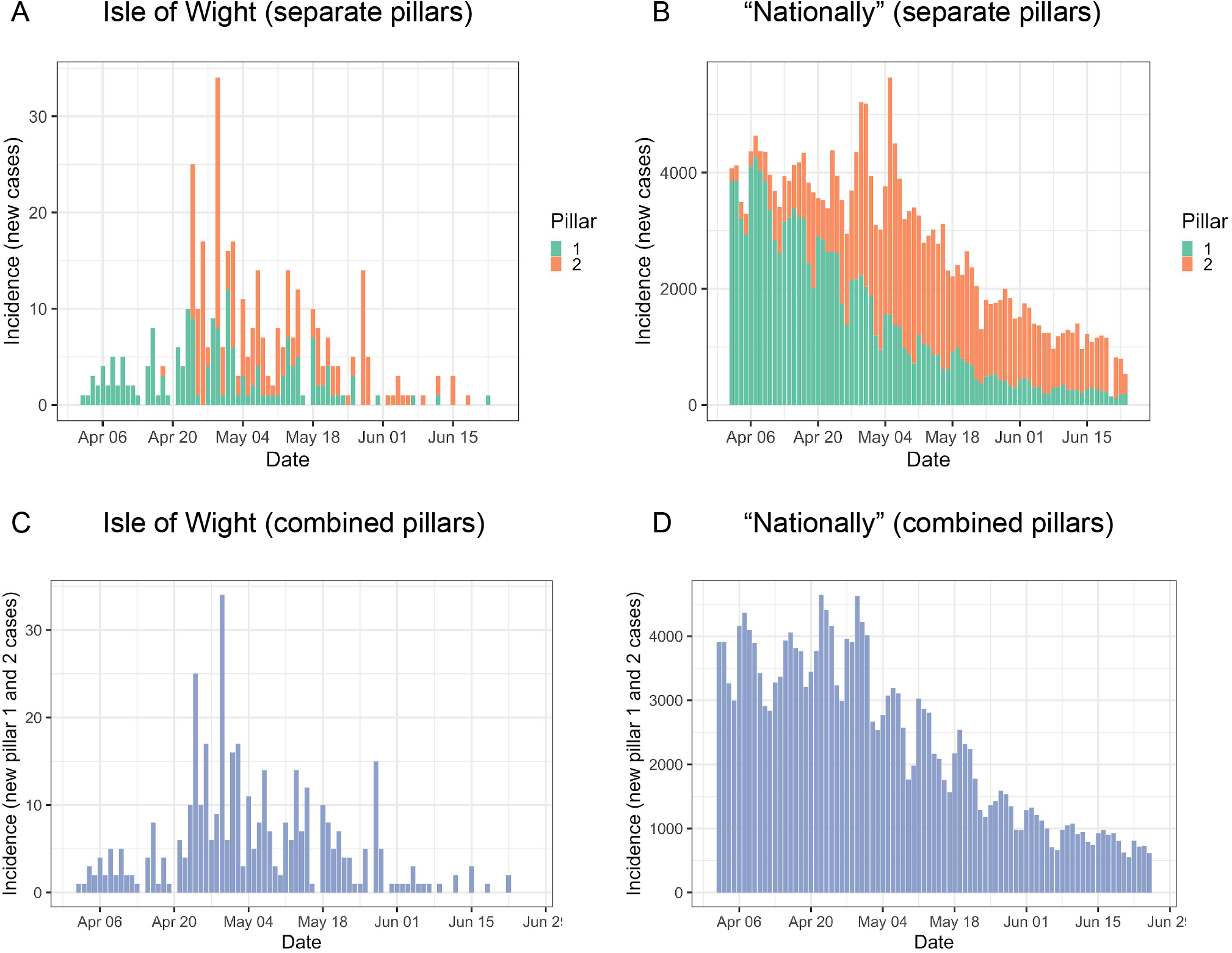
Daily incidence (A) on the Isle of Wight and (B) “nationally” (England for Pillar 1, UK except Wales for Pillar 2). (C) and (D) use the newly-released “combined” pillars data where Pillar 2 has been deduplicated. Each of these provides only a rough approximation of the true incidence because cases are recorded by the date the specimen was taken, not the time the individual became infected, and tests are expected to be performed at different stages of infection between the two pillars.

### Incidence decreased after the TTI intervention on the Isle of Wight more sharply than the national trend

To compare between UTLAs with accurate timing information we used Pillar 1 daily case data on the Isle of Wight (Figure 3A, green) and inferred the incidence of new infections (Figure 3A, red). The number of new infections each day on the island was generally decreasing from mid-April, although we do not have the data to be able to deduce whether this was entirely the impact of the lockdown or whether the result is exaggerated by lack of test availability. We hypothesise that the apparent uptick in new infections in late April is largely an artefact of the increased availability of tests after the TTI intervention. We see a particular decline in incidence straight after the launch of the TTI intervention on 5 May (dashed line). Since the back-calculation relies on using around 10 days of future data, the inferred incidence over the final 10 days is increasingly prone to underestimation and is shown shaded in grey in Figure 3A. In Figure 3B and Figures 4–6 we truncate our results on 14 June for this reason. The trend in incidence looks encouraging for the Isle of Wight but it is important to compare the incidence with other areas in England that did not see the launch of the TTI programme on 5 May. Figure 3B shows the inferred incidence curves scaled per capita for each of the UTLAs, with the Isle of Wight shown in red. We see that in mid-April per capita incidence on the Isle of Wight was around average for English UTLAs, but by late May the incidence on the Isle of Wight was very low. COVID-19 incidence therefore decreased more sharply on the Isle of Wight than in other areas of England.

**Figure 3:**
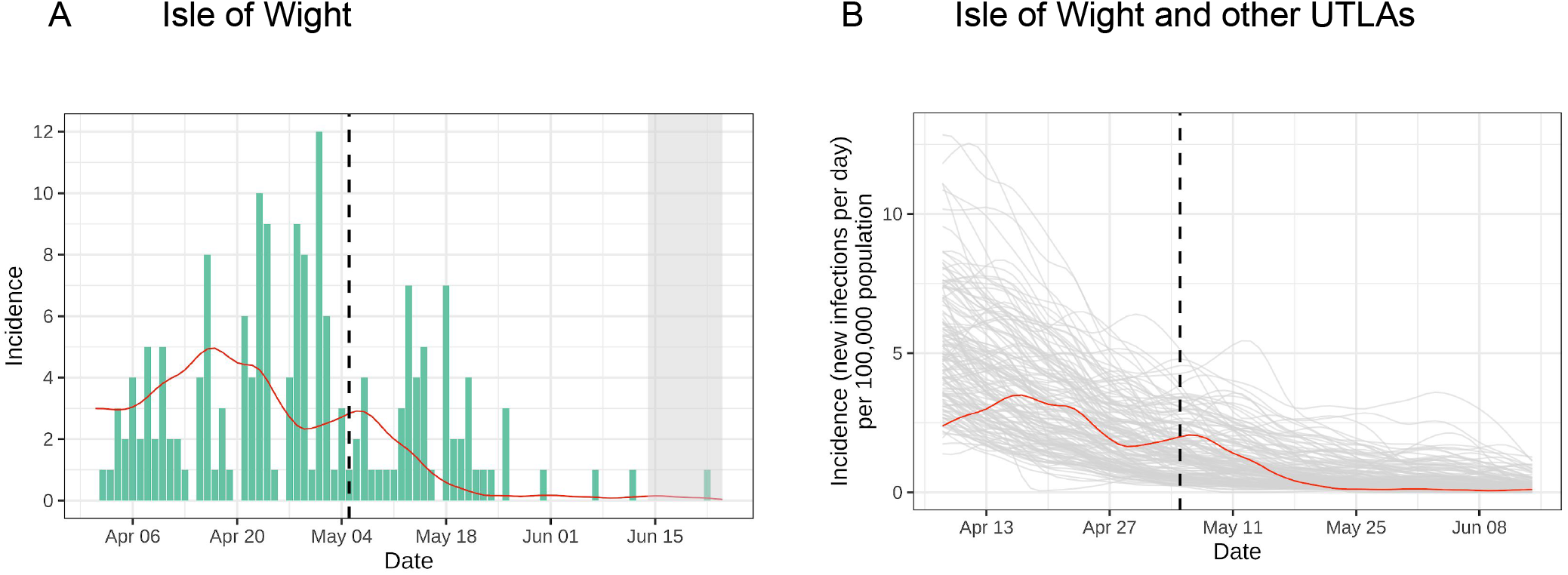
Inferred incidence using Pillar 1 data. (A) back-calculated estimation of incidence of new infections (red) from case data (green); (B) back-calculated incidence of new infections per capita for each UTLA, with the Isle of Wight in red.

**Figure 4:**
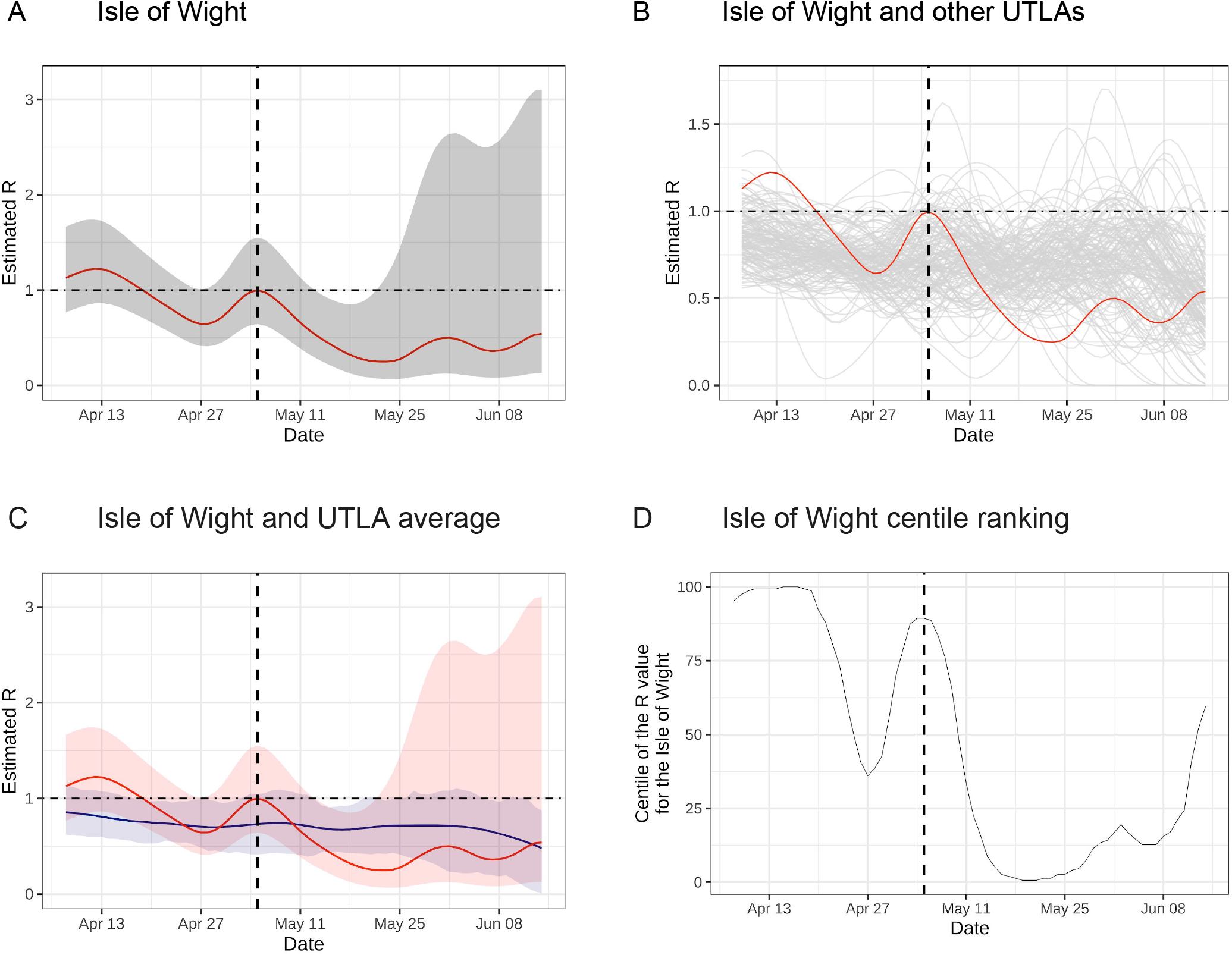
Estimated R using Pillar 1 data. (A) estimated R on the Isle of Wight (red) with confidence intervals shown in grey; (B) estimated R on the Isle of Wight (red) and other UTLAs; (C) estimated R on the Isle of Wight with credibility intervals (red), and average R across other UTLAs with 5%-95% range (blue); (D) estimated R on the Isle of Wight as a centile of other UTLAs.

### R decreased after the TTI intervention on the Isle of Wight more sharply than the national trend

The value of the reproductive number R over time is shown in Figure 4. In these figures we report for each date the estimate of R for the week which starts 3 days before and ends 3 days afterwards. On the Isle of Wight, R declined rapidly after the TTI programme’s introduction (dashed line) from a value of 1.0 on 5 May to 0.25 on 23 May. It then followed a fluctuating and gradually rising trend to 0.54 on 14 June. However, incidence was very low during the period 23 May - 14 June (Figure 3A) so the confidence intervals on R are very wide (Figure 4A, grey). In Figure 4B we plot the Isle of Wight’s time-varying reproduction number R alongside those of the other UTLAs. The decline from 5 May to 23 May was much sharper than that of a typical UTLA. In Figure 4C we compare R on the Isle of Wight (red) with the average R of other UTLAs and in Figure 4D as a centile of all UTLAs. On 5 May the reproduction number R on the Isle of Wight was high compared to the national average, shown in Figure 4C by the red line nearing the top of the blue 95% range and in Figure 4D ranked in the 89th centile. By 23 May it was well below the 5% range (Figure 4C, blue) and ranked lowest of all UTLAs 19-21 May (Figure 4D). R therefore decreased more sharply on the Isle of Wight than in other areas of England.

In the final few weeks of the study an increasing majority of cases were recorded as Pillar 2 (Figure 2B), and Pillar 1 case numbers in all UTLAs decreased to nearly zero. We do not know how consistent the magnitude of this shift from Pillar 1 to Pillar 2 case numbers was across UTLAs. The effect of the shift is that incidence and R calculated only from Pillar 1 decreased rapidly to zero for all UTLAs (Figures 3B and 4C). The centile ranking of the Isle of Wight (Figure 4D) therefore became higher but less meaningful (the statistical test for deviation loses power).

### Nowcasting

The “nowcast” for each UTLA (Figure 5A) shows that throughout April the epidemic on the Isle of Wight had a size and growth comparable to that of an average UTLA in England, between the 25th and 75th centiles (Figure 5B). Following the TTI intervention the prospects for the Isle of Wight looked significantly better than those for the majority of UTLAs, reaching the second centile in late May. In the final few weeks of the study, incidence and R were both very low leading to stochasticity and thus ceasing to be useful measures to compare epidemic trends across areas.

**Figure 5:**
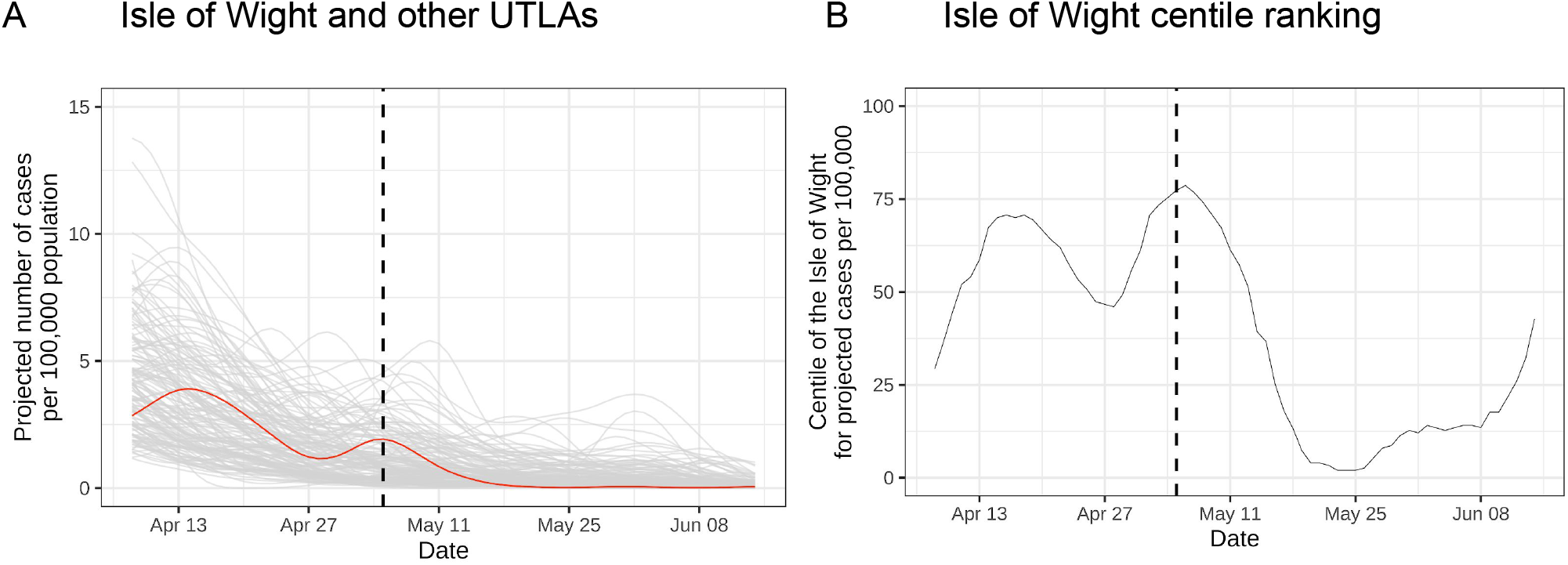
Simple “nowcast” to quantify the combined effects of R and incidence using Pillar 1 data. (A) nowcast for each UTLA, with the Isle of Wight’s line in red, and (B) centile of the Isle of Wight’s projection ranked against the other areas.

Our results comparing incidence, R and nowcasts across UTLAs using Pillar 1 data (Figures 3B, 4B and 5A) are available in the *EpiNow-C19* web application [1] where they can be explored in more detail. These are provided alongside a daily extension of the analyses using the available combined pillars dataset, noting that this data provides a better estimate of the number of infections than Pillar 1 data alone but presents challenges for estimating timings and R.

### The Isle of Wight has a lower R compared to the rest of England after the intervention using a synthetic control approach

Since the demographics and trajectories of the sub-epidemics vary across UTLAs we used a ‘synthetic control’ approach [20] to create a more comparable control group for the Isle of Wight. We combined data from all other English UTLAs and weighted their contributions to match the evolution of R on the Isle of Wight before the introduction of the TTI pilot (Figure 6, left hand panels). The choice of matching variables is to some extent subjective and can influence the result of the analysis. We therefore consider two different scenarios, first using weekly averages of the estimates of R alone, and second using additional demographic information. In both cases we use a cross validation approach to guard against overfitting. Table S1 shows the weights on the matching variables for the two scenarios. When we add population density and measures of deprivation to Scenario 2, almost no weight is allocated to these variables and the overall results remain unchanged (data not shown).

**Figure 6:**
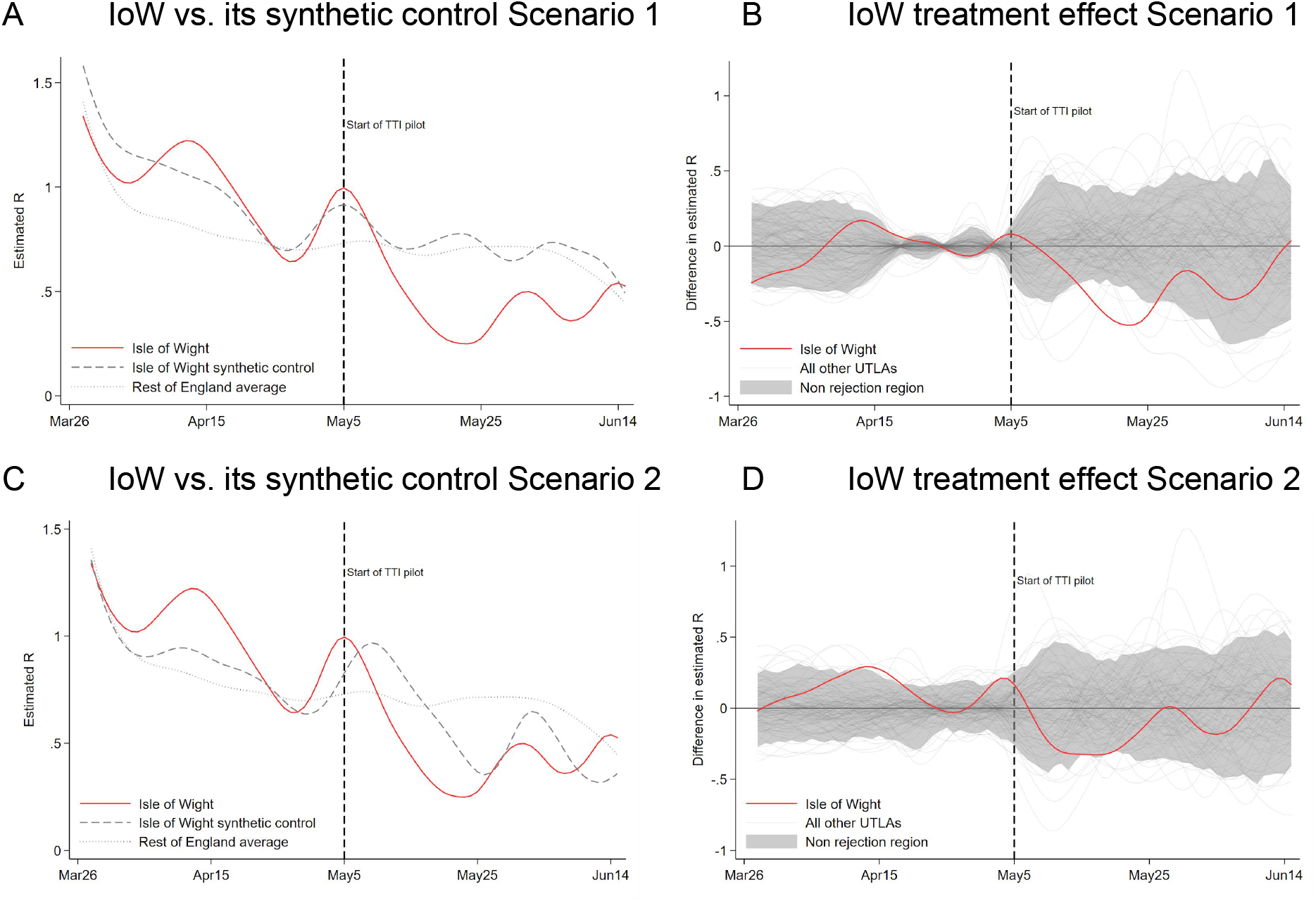
Comparison between R on the Isle of Wight and the other areas in England using a synthetic control approach and Pillar 1 data. (A) Synthetic control using the average R in each of the three weeks of the training/validation period as matching variables. This scenario gives 71% weight to the East Ridings of Yorkshire, 28% to Doncaster and 1% to Sandwell in constructing the synthetic control. (B) Difference between each UTLA and its respective synthetic control (Scenario 1). The grey area contains 95% of all UTLAs. A line outside the area is classified as significantly different from zero at the 5% level. (C) Synthetic control using the average R in each of the three weeks and age and ethnicity variables (proportion of the population aged 0-19, 20-44, 45-64, 65-74 and 75+; the proportion of the population being white, Asian, Black and African Caribbean) as matching variables. This approach gives 64% weight to Dorset, 32% to North Somerset, 3% to the East Ridings of Yorkshire and 1% to Kensington and Chelsea in constructing the synthetic control. (D) Difference between each UTLA and its respective synthetic control (Scenario 2).

Both synthetic control scenarios show that R is lower for the Isle of Wight than for its synthetic control during almost the entire period after the introduction of the TTI pilot. To establish the statistical significance of this result we perform a permutation test by constructing synthetic controls for all other UTLAs in England. We compute the difference between each UTLA and its synthetic control (Figure 6, right hand panels). Using this method we find that the difference for the IoW is statistically significant for part of May in both scenarios at the 5% level (see Supplementary Material for more details).

### Combining Pillar 1 and Pillar 2 test data: The Isle of Wight saw a significant decrease in R after the TTI intervention compared to the rest of England

We used three different datasets (Pillar 1, Pillar 2 and the combined data) to obtain Maximum-Likelihood estimates of R after the TTI interventions. Figure 7A shows the results across the different pillar datasets. The reproduction number R on the Isle of Wight was consistently lower than the “national” rate, by which we mean England for Pillar 1 and the combined dataset, and the UK except Wales for Pillar 2 data. The national estimates have tight confidence intervals (Figure 7A, error bars) thanks to being informed and constrained by more data. R was lower for the Isle of Wight than nationally for data from Pillar 1 (*p* = 6 × 10^−6^, likelihood ratio test), from Pillar 2 (*p* = 5 × 10^−4^) and from both combined (*p* = 2 × 10^−11^). We also used Pillar 1 data to compare R on the Isle of Wight to R for each UTLA individually before the intervention from 1 March (Figure 7B) and afterwards (Figure 7C). By this estimation the Isle of Wight experienced a striking reduction in its reproduction number, going from 1.3 in the period before the TTI intervention to 0.5 in the study period afterwards. This is a particularly notable change when compared with other UTLAs as it went from having the third highest reproduction number before the intervention to the tenth lowest afterwards.

**Figure 7:**
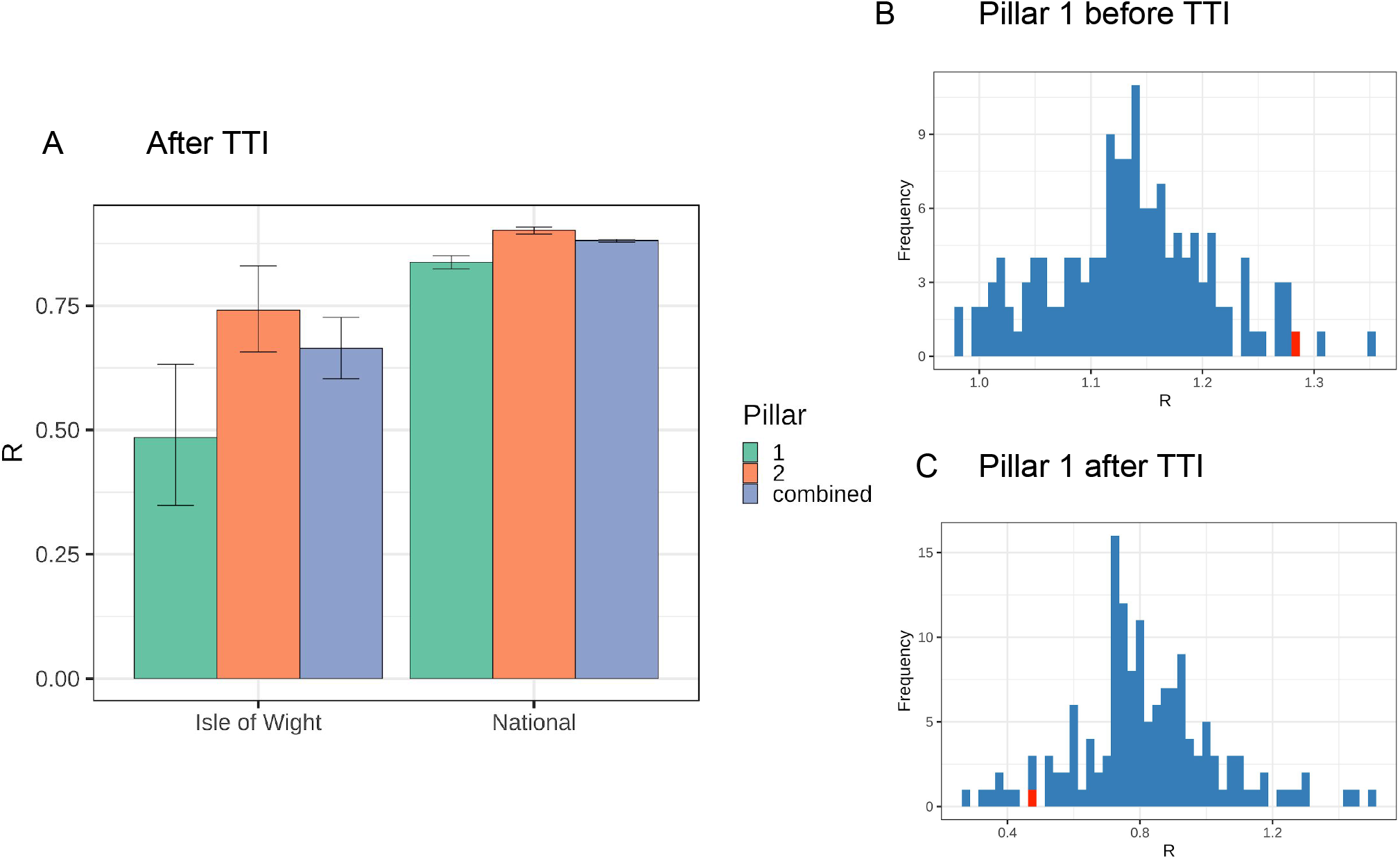
Maximum-Likelihood estimates of R: (A) on the Isle of Wight and nationally, for each data set; (B) for each UTLA using Pillar 1 data, with the Isle of Wight in red, before the TTI intervention; (C) for each UTLA using Pillar 1 data, with the Isle of Wight in red, after the TTI intervention.

## Discussion

We analysed the course of the COVID-19 epidemic on the Isle of Wight before and after the launch of the Test, Trace, Isolate (TTI) programme including a test version of the mobile phone contact tracing app and compared it to the epidemic in other areas of England and the UK. We focussed on total incidence, per capita incidence, and the effective reproduction number R and compared each using different approaches. We used an improved method to calculate R by taking into account variable lag times between infection and case counts; the method can be generalised to account for different distributions of lag times across data sources.

All analyses paint a consistent and encouraging picture. In mid-April the Isle of Wight was experiencing moderate-to-high incidence per capita and a higher reproduction number R than a typical UTLA. The launch of TTI on the Isle of Wight was followed by a short phase in which case numbers increased, as expected with increased testing. However, the inferred incidence of new infections and the reproduction number R declined sharply to below the English average, and below predicted levels for a synthetic control, immediately upon introduction of TTI. This finding is consistent across our analyses of Pillar 1, Pillar 2 and the combined pillars datasets, and in both our Bayesian and Maximum-Likelihood analyses.

These results are correlative: rapid decreases in the rate of new infections on the Isle of Wight could be explained by multiple causes. Comparisons are statistically significant for every way in which we analysed the data, suggesting that the effect of chance fluctuations alone is unlikely. The introduction of TTI was accompanied by community discussions and national publicity. The success could be due to the attention that the start of the programme on the Isle of Wight received leading to increased care and physical distancing, although changes in movement are not apparent from Google mobility data [23] (Supplementary Figure S1). All inhabitants received a letter and an invitation to download the app and contact tracers might have been highly motivated to make the first phase of the launch a success. The start of the programme and the testing of the app might have led to a positive community response and an increased awareness. A hospital-based app usability study concluded that the app had been embraced and adopted well during the initial trial period, whilst making recommendations for improvements for future versions [24]. Contact tracing could have contributed to the drop in infections, both through the manual/CTAS system, and through the use of the app by 38% of inhabitants. Alternatively, the success in reducing infections on the Isle of Wight might be attributable to reasons other than the launch of the TTI programme.

The national TTI programme was launched on 28 May during a period of gradual relaxation of lockdown measures which began on 10 May. It is possible that this is why we did not see a strong decrease in incidence and R at this point; it was also towards the end of our period of study so there was little opportunity to observe a more gradual decline.

This study has limitations. Data from the Contact Tracing and Advisory Service (CTAS) and time series of cases traced by the app are not yet publicly available. This study is therefore unable to evaluate the effect of the app independently of the effect of the wider TTI programme. We did not adjust for changes in testing practice, and so likely overestimated the reproduction number over time when widespread community testing became available in the week beginning 5th May in the Isle of Wight, and in other areas in the week beginning 18 May. We hypothesise that these biases become less pronounced after one week. The Isle of Wight’s synthetic controls are not perfect matches, as is evident from several time periods that are significantly different between the Isle of Wight and its synthetic control before the start of the TTI pilot. This is, however, not surprising given that the Isle of Wight’s unusual epidemic trajectory and geography make it harder to find a perfect synthetic control.

We are currently using the Public Health England data which is published daily [2] to provide an approximate surveillance and nowcasting application *EpiNow-C19* [1]. To improve the accuracy of this and to build on the results of this study, we recommend that local area data is made publicly available for all regions of the UK, either separated by Pillar or with similarly informative timing information such as the date of onset of symptoms. We encourage further analyses comparing sub-epidemics across geographical areas as the TTI programme evolves, with focused data collection that allows evaluations of the separate TTI components. In particular, knowing the proportion of cases from individuals who were already quarantined at the point of diagnosis is a strong indicator of the effectiveness of TTI. Tracking the progression of local sub-epidemics and identifying the key determinants of successful suppression will be crucial for informing local and national non-pharmaceutical intervention strategies.

## Data Availability

Most of the data is publicly available at the links below. We obtained "Pillar 2" data for the Isle of Wight via private correspondence with Public Health England; we explain in the text how this can be almost exactly deduced from the publicly available data.

https://coronavirus.data.gov.uk/#category=nations&map=rate

https://github.com/tomwhite/covid-19-uk-data

## Supplementary material

### Supplementary Methods

#### Data on incidence of confirmed cases by area

We used publicly available reports of Pillar 1 and Pillar 2 data until 30 June 2020 [9,10]. Pillar 1 data was available by Upper Tier Local Authority (UTLA) whereas Pillar 2 was reported as a national daily total. On 1 July 2020, Public Health England stopped reporting Pillar 1 cases alone, and on 2 July after a deduplication procedure they began publishing daily Pillar 1 and 2 case numbers combined at the levels of countries, regions, UTLAs and Lower Tier Local Authorities (LTLAs) [25,26]. Although Pillar 1 data does not give a complete picture of confirmed cases and is restricted to the UTLA level, we focus on Pillar 1 cases for our estimation of incidence and the time changing reproduction number R(t) because these are the most consistently reported over time and across areas. The combined pillars dataset presents challenges for interpreting incidence of new infections because of differences in timing: cases are reported by date of swab test, which for a Pillar 2 test is likely to be shortly after symptom onset so around 4-7 days post-infection, but for Pillar 1 is likely to be shortly after admission to hospital so around 7-12 days post-infection [12]. Pillar 2 daily case data for the Isle of Wight was obtained via private communication with Public Health England. This data can be approximated by subtracting the Pillar 1 data from the combined data; the differences caused by the deduplication procedure are slight as can be seen by comparing Figures 2A and 2C.

#### Back-calculation of infection times

For each of the 150 areas of England under study, we have a time-series of reported cases *C*_*t*_, where *C*_*t*_ is the number of cases newly diagnosed via hospital testing, where the swab was taken on day *t*. We used a back-calculation approach to infer the time-series of infection times of these cases *I*_*t*_. Let ζ_*s*_ be the probability distribution of times elapsed *s* between being infected and being swab-tested in hospital. For a case infected at time *t-s* the probability that they are swab-tested at time *t* is ζ_*s*_. Conversely, for a case swab-tested at time *t*, the probability that they were infected at time *t-s* is 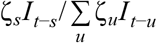, i.e. the backwards-in-time delay distribution is not in general equal to the forwards-in-time distribution. However, we make the simplifying assumption that incidence changes sufficiently slowly across the range of ζ_*s*_ (about a week) that 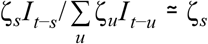, and so we estimate that, of the people who became Pillar 1 confirmed cases, the number infected at time *t* is 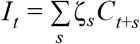.

For ζ_*s*_ we took the convolution of the incubation period distribution and the distribution for time from symptoms to hospital visit, making the assumption that the majority of the Pillar 1 tests were for patients. For the incubation period we used meta-analysis results indicating a lognormal distribution with mean 5.42 days and standard deviation 2.7 days [14] (specifically, the results excluding those of Ma et al for which bias was detected [14]). For time from symptoms to hospital visit time we used a gamma distribution with mean 5.14 days and standard deviation 4.2 days [12].

Back-calculation of incidence leads to right-censoring towards the present by an amount corresponding to roughly the mean of ζ_*s*_, and so we truncate analyses 10 days before the last date of “reliable” data. That is, we truncate analyses at 14 June because accurate estimates after this time would require complete data beyond 24 June, but case numbers within the last 5 days of the dataset (25-29 June) should be considered incomplete [9].

### Renewal equation estimation for R(t)

The time-varying effective reproduction number *R*(*t*) is a standardized measure for the rate of spread of an infection at time t. Several definitions exist; here we used the instantaneous reproduction number [17]. Use of this measure and back-calculation from reporting dates were recently recommended by a survey of methods [27] and has been implemented across many different countries for COVID-19 [28]. A Bayesian method of estimation of this measure is implemented in the software package EpiEstim [18,19]. Specifically, our definition of *R*(*t*) was 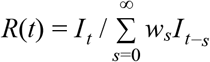 where *I*_*t*_ is the number of inferred infections on day *t* and *w*_*s*_ is the generation time distribution for day *s*. If the Bayesian prior for *R*(*t*) is a gamma distribution with shape and scale parameters (*a, b*) and if the number of infections is Poisson distributed, then the posterior distribution for the reproduction number is also gamma distributed [18] with parameters (*a*′, *b*′) given by 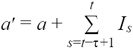 and 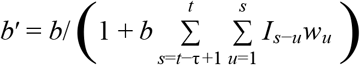.

To reduce oversensitivity to the priors (*a, b*) when the number of infections is very low, instead of reporting as the central estimate of *R*(*t*) the posterior mean (which is *a*′*b*′ and is the default in EpiEstim) we report instead, as the central estimate, the posterior mode, namely (*a*′ − 1)*b*′. The choice of posterior mode as a central tendency is common in Bayesian statistics, and reduces the tendency to obtain high estimates of *R*(*t*) when there are very few reported cases in an area. We report the usual 95% credibility intervals.

### Nowcasting

The future course of an epidemic depends on both the current number of infectious individuals and the effective reproduction number R. In the last month of this study the Isle of Wight had a consistent pattern of recording just one Pillar 1 case every 5-7 days. Although we know that there were also Pillar 2 cases, the Pillar 1 case count is consistent with a single chain of transmission where one individual infects exactly one other: a reproduction number of 1.

However, the short-term forecast is clearly better for the island than it would be for another UTLA with a reproduction number of 1 but many Pillar 1 cases per 100,000 population. We combine R with incidence per capita to provide a simple “nowcast” for each UTLA in the following way: for each UTLA on each date we consider the week which starts 3 days before and ends 3 days afterwards. We multiply the estimated value of R in that week by the mean incidence per capita for that week to provide a simple projection of the expected number of new cases per capita per day in that area in the near future.

### Creating synthetic controls for the 150 UTLAs in England

The ‘synthetic control’ method was developed to evaluate the effect of a treatment on the aggregate level, e.g., regions, when only one region is treated [20, 21]. The method provides a data-driven manner of constructing a more similar comparison unit. The synthetic control is constructed as a weighted average of all ‘donor’ (control, untreated) units. The vector *W* of donor unit weights is chosen based on matching variables *X* such that the outcome variable of the synthetic control most closely resembles the outcome variable of the treated unit in the pre-treatment period. In our setting, the Isle of Wight is the treated region and the treatment is the introduction of the TTI pilot. The donor units are all other UTLAs in England. The outcome variable *Y* is the daily value of R, estimated as described above. As matching variables *X* we use age bands, ethnicity shares in the population, and weekly averages of the estimated values of R.

We used the *synth* package in Stata to construct the synthetic control. The weights are chosen over *T* days and *N = 149* donor units. Define *Y*_treat_ as a (*T* × 1) vector of the treatment unit’s outcome variable, i.e., R on the Isle of Wight. *Y*_donor_ is a (*T* × *N*) matrix of the donor units’ outcome variable. There are J matching variables. Define *X*_treat_ = (*X*_treat_ ^1^, …, *X*_treat_ ^*J*^)’ as a (*J* × 1) vector of matching variables for the treatment unit and similarly *X*_donor_ as the corresponding (*J* × *N*) matrix of matching variables for the donor units. Define *W* as an (*N* × 1) vector of weights that sum to one: *W* is a possible *synthetic control*. Finally, define *V* as a (*J* × *J*) diagonal matrix of matching variable weights. Thus *W* contains the weights across donor units and *V* contains the weights across matching variables. Then the optimal synthetic control W* and the associated symmetric matrix V* solve the following minimisation problems:

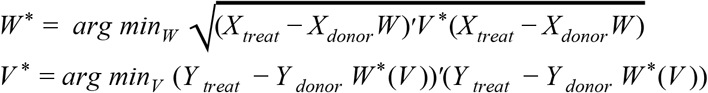

To provide robustness against overfitting, we determined the matching variables weights (*V*) by cross validation. Specifically, we split the pre-treatment period into two equal-length periods, the *training period* and the *validation period*, so *T* = 19 days for each of the two periods. The minimisation was conducted twice, first to find a *W* ^*train*^ using training period data and a *V* ^*val*^ using validation period data, and again using data from the validation period to find *W* ^*^ by setting *V* ^*^ = *V* ^*val*^ found in the first minimisation.

To test whether the difference between the Isle of Wight data and its synthetic control is statistically different from zero, we used a permutation test. First, we created synthetic controls for all 150 areas in England and calculated the difference between the outcome variable in each area and its synthetic control. The Isle of Wight was deemed significantly different from its synthetic control at the 5% level if it was in the 2.5 centile of the areas with the largest negative or the 2.5 centile of the areas with the largest positive difference to their respective synthetic control.

**Table S1:** Matching variable weights for the Isle of Wight synthetic controls in different scenarios

| Matching variable | Scenario 1 | Scenario 2 |
| --- | --- | --- |
| Average R in the first five days of the training or validation period | 8% | 0% |
| Average R in week 2 of the training or validation period | 37% | 0.001% |
| Average R in week 3 of the training or validation period | 55% | .001% |
| Fraction white |  | 64% |
| Fraction Asian / Asian British |  | 24% |
| Fraction Black / African / Caribbean / Black British |  | 8% |
| Fraction mixed / multiple ethnic groups |  | 0.7% |
| Fraction other ethnic group |  | 0.7% |
| Fraction 0-19 years |  | 0.2% |
| Fraction 20-44 years |  | 1.3% |
| Fraction 45-64 years |  | 0.3% |
| Fraction 65-74 years |  | 0.3% |
| Fraction 75+ years |  | 0.04% |

### Maximum-Likelihood estimation of R

We estimated R(t) for Pillar 1 data because it is comparable across time and space; however, valuable information is present in Pillar 2 data based on community testing. Because this was not collected consistently over time, and community testing was introduced on the Isle of Wight before the rest of the country (Figure 1), we performed three separate analyses using Pillar 1 data, Pillar 2 data and the combined dataset, estimating for each the exponential rate of change *r* over defined time periods of analysis that clearly pre-date and post-date the introduction of TTI. For a given series of cases *C*_*t*_ in a time period [*t*_1_, *t*_2_], we estimated the rate of change by maximising a simple Poisson log likelihood 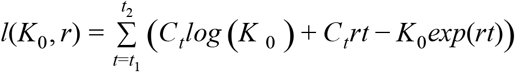 with respect to the intercept *K*_0_ and the slope *r*. We used the likelihood ratio test to compare the slope between two time series (that for the Isle of Wight versus that for elsewhere): we compared the likelihoods obtained from a model constrained to use the same slope *r* for the two series versus a model with freedom to select a different slope for each series. Ranges [*t*_1_, *t*_2_] were chosen so as to represent the time period of interest, taking into account delays in reporting of cases relative to underlying infections, namely 5 days for Pillar 2 cases and the combined data, and 10 days for Pillar 1 cases alone. We also remove the final 5 days of data available for each data set since these may be incomplete [9]. The start and end dates are given explicitly in Table S2. Finally, we use the relation 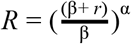 to determine R from r [22] when the generation time is gamma-distributed, using values α = 6.6 and β = 1.2 [Ferretti et al.(in prep)].

**Table S2:** start and end dates [t_1_, t_2_] used in our Maximum-Likelihood estimation of the growth rate r.

| | $t_1$ Isle of Wight | $t_1$ National | $t_2$ |
| --- | --- | --- | --- |
| Pillar 1 | 15 May | 28 May | 24 June |
| Pillar 2 | 10 May | 23 May | 19 June |
| Combined | 10 May | 23 May | 26 June |

**Figure S1:**
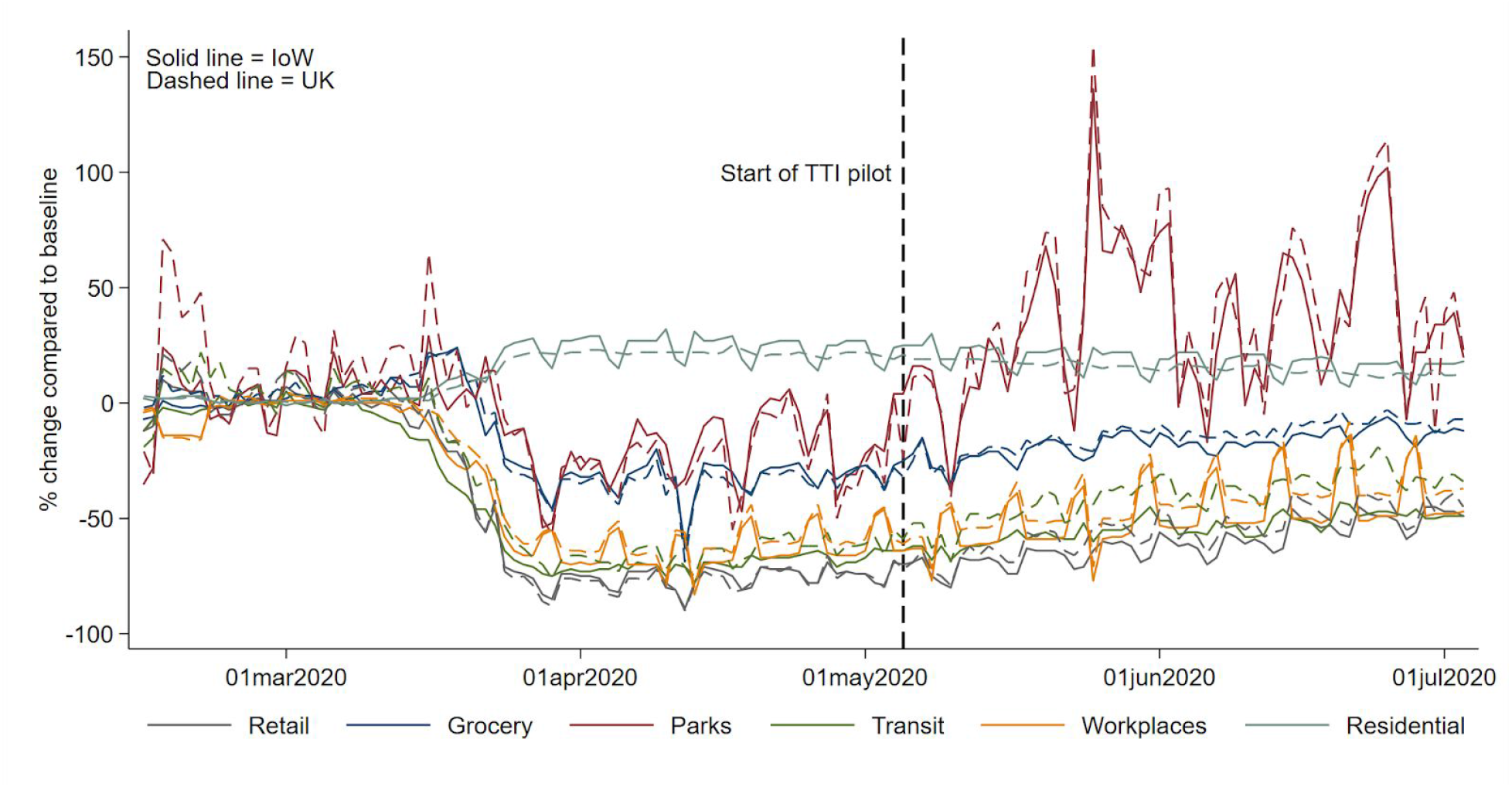
Google mobility data shows similar trends for the Isle of Wight (solid) and the UK (dashed) over the periods of lockdown and easing of restrictions.

